# Characteristics of Cerebellar Cognitive Affective Syndrome (CCAS) in patients with acute cerebellar stroke and its impact on outcome

**DOI:** 10.1101/2023.08.03.23293529

**Authors:** Tokuaki Shinya, Yamauchi Kota, Shota Tanaka, Kei Goto, Shuji Arakawa

## Abstract

**Background:** Recently, cerebellar cognitive affective syndrome-scale (CCAS-S) was developed to assess CCAS—a syndrome in which cognitive impairment is caused by cerebellar lesions. Literature regarding CCAS in patients with stroke is scarce, and its impact on outcomes is unclear. This study aimed to evaluate CCAS in patients with acute cerebellar stroke and examine its relationship with the outcomes.

**Methods:** Patients who experienced acute cerebellar stroke for the first time and were hospitalized in Steel Memorial Yawata Hospital within 7 days of stroke onset between April 2021 and April 2023 were included in this observational study. CCAS-S, Mini-Mental State Examination (MMSE), and Scale for the Assessment and Rating of Ataxia (SARA) scores were measured 1 week after stroke onset, and Functional Independence measure (FIM)/Barthel Index (BI) at discharge, physical function, activities of daily life (ADL), length of hospital stay, and outcome (discharge destination) were evaluated. The Mann–Whitney U test was used to compare CCAS-S scores and variables.

**Results:** Thirteen consecutive patients with acute cerebellar stroke (9 women) and age-and sex-matched healthy controls (7 women) were included. The MMSE score was within the normal range in all patients; however, patients with stroke had a lower total CCAS-S score (median 72, interquartile range [IQR] 66–80) and a higher number of failed tests (median 4, IQR 3–5) than those of healthy controls. Significant deficits were observed in semantic fluency (p=0.008), category switching (p=0001), and similarity (p=009). Possible, probable, and definite CCAS were diagnosed in 2, 1, and 10 patients, respectively. Patients discharged home showed better SARA and FIM/BI scores but similar CCAS-S scores compared to those discharged to rehabilitation hospitals.

**Conclusion:** CCAS along with impaired executive and language functions is frequently observed in patients with acute cerebellar stroke; however, impaired motor function, and not CCAS, influences the outcome.

## Introduction

The cerebellum influences motor functions, such as walk, balance, and coordinated movement; however, until recently, its function in language, cognition, and affect has not received much attention [1]. In the 1980s, Schmahmann [2] defined a new clinical symptom, cerebellar cognitive-affective syndrome (CCAS), a cognitive impairment as a result of damage to the cerebellum. CCAS is characterized by four domains: executive dysfunction, visuospatial cognitive impairment, language dysfunction, and affective disorders. The connection between the cerebellum and cognition has become clearer with advances in research related to anatomical aspects of CCAS, functional imaging of healthy individuals, and neuropsychological aspects of patients with cerebellar lesions. In stroke patients, the incidences of cognitive impairment and personality disorders are high at 86% and 64%, respectively [3]. However, owing to the low awareness of CCAS in clinical practice, mild cognitive dysfunction is often overlooked [4]. This can be attributed to the absence of a strict definition or diagnostic model for CCAS, leaving the diagnosis at the clinician’s discretion, usually based on a comprehensive neuropsychological examination. In addition, the wide variety of CCAS symptoms require extensive and time-consuming assessment. Many studies have identified efficient assessment methods, however, they vary across studies and are inconsistent [5–7]. In 2018, Hoche et al. [8] developed the CCAS Scale (CCAS-S), a screening test for CCAS, that consists of 10 neuropsychological items and requires 12–15 min for assessment. In recent years, the number of studies using the CCAS-S has increased, but most have been conducted in patients with spinocerebellar ataxia or Friedreich’s ataxia (FRDA) [9, 10]. In contrast, very few studies utilizing the CCAS-S in acute stroke patients have been conducted, and only one has investigated the prevalence of CCAS and utility of CCAS-S in stroke patients [11]. Therefore, this study aimed to investigate the characteristics of CCAS in acute cerebellar stroke patients using the CCAS-S. In addition, we investigated relationship between the CCAS-S and physical function, activities of daily living (ADL), length of hospital stay, and outcome.

## Methods

### Patients

In this study, acute stroke patients who were hospitalized at Steel Memorial Yawata Hospital, Japan within seven days of stroke onset between April 2021 and April 2023 and met the following criteria: (1) first-ever stroke; (2) computed tomography (CT) or magnetic resonance imaging (MRI) showing cerebral infarction or hemorrhage confined to the cerebellum; and (3) independent ADL without dementia before stroke onset were included. The exclusion criteria were: (1) lesions other than those in the cerebellum and (2) a history of neurological disease or mental disorder. Age-and sex-matched healthy patients with independent ADL and normal cognitive function were recruited as controls.

### Ethical Approval

In accordance with the Declaration of Helsinki, this study was accepted by the Ethics Review Committee of Steel Memorial Yawata Hospital (approval number 20-62-02), and careful consideration was given to the handling of personal information. In addition, written informed consent was obtained from all patients before their participation in the study.

### Statements and Declarations

The study report was organized according to the Strengthening the Reporting of Observational Studies in Epidemiology (STROBE) statement [12].

### Assessment

The CCAS-S, Mini-Mental State Examination (MMSE), and Scale for the Assessment and Rating of Ataxia (SARA) were measured a week after stroke onset. Additionally, Functional Independence Measure (FIM) and Barthel Index (BI) at discharge, length of hospital stay, and outcome (discharge destination: home or rehabilitation hospital) were evaluated.

The CCAS-S consists of 10 test items scored from 0 to 120 points, with a cutoff value of 82 points [8]. For each item, a cutoff value was established, and possible (sensitivity, 95%; specificity, 78%), probable (sensitivity, 82%; specificity, 93%), and definite CCAS (sensitivity, 46%; specificity, 100%) were defined as one, two, and three or more failed items, respectively. The SARA consists of eight items, ranging from 0 for no ataxia to 40 for severe ataxia [13] and is useful for assessing the severity of stroke-related ataxia [14]. Each item was assessed as previously described [8, 13].

### Statistical Analysis

Data are presented as median with interquartile range (IQR) or as frequencies and percentages. The Mann–Whitney U test was used to compare CCAS-S scores and other variables between with acute cerebellar stroke patients and healthy controls. For the analyses of respective CCAS-S item, in addition to the original cutoff value, the first quartile of the healthy controls was used as the cutoff value as our patients were older than those in the original CCAS-S study [8]. Additionally, we compared these variables between patients discharged to their home and those discharged to a rehabilitation entre. SPSS Statistics (version 23, IBM, Armonk, NY, USA) was used for statistical analysis. A p<0.05 was considered statistically significant.

## Results

During the research period, 301 acute stroke patients were admitted to our hospital, 14 of whom met the inclusion criteria. After excluding one patient as their assessment was impeded due to comorbid acute myocardial infarction, thirteen cases were included in the final analysis. All the patients underwent rehabilitation during hospitalization.

Patient characteristics of both the study groups are shown in Table 1. The median age was 75 years (IQR:73–79). Cerebellar lesions were ischemic in seven, hemorrhagic in six, left-sided in five, right-sided in seven, and bilateral in one patient. The affected vascular territories were the superior cerebellar artery (SCA) in 8 cases, the anterior inferior cerebellar artery (AICA) in 1 case, and the posterior inferior cerebellar artery (PICA) in 4 cases. A week after the onset, the median CCAS-S, MMSE, and SARA score was 72 (IQR: 66–80), 28 (IQR: 26–29), 6 (IQR: 2–15), respectively. At discharge, the median FIM and BI was 96 (IQR: 85–113) and 80 (IQR: 70–100), respectively. The median hospitalization days was 18 days (IQR: 13–21). Nine patients were directly discharged from the hospital and four were transferred to a rehabilitation centre.

**Table 1.**
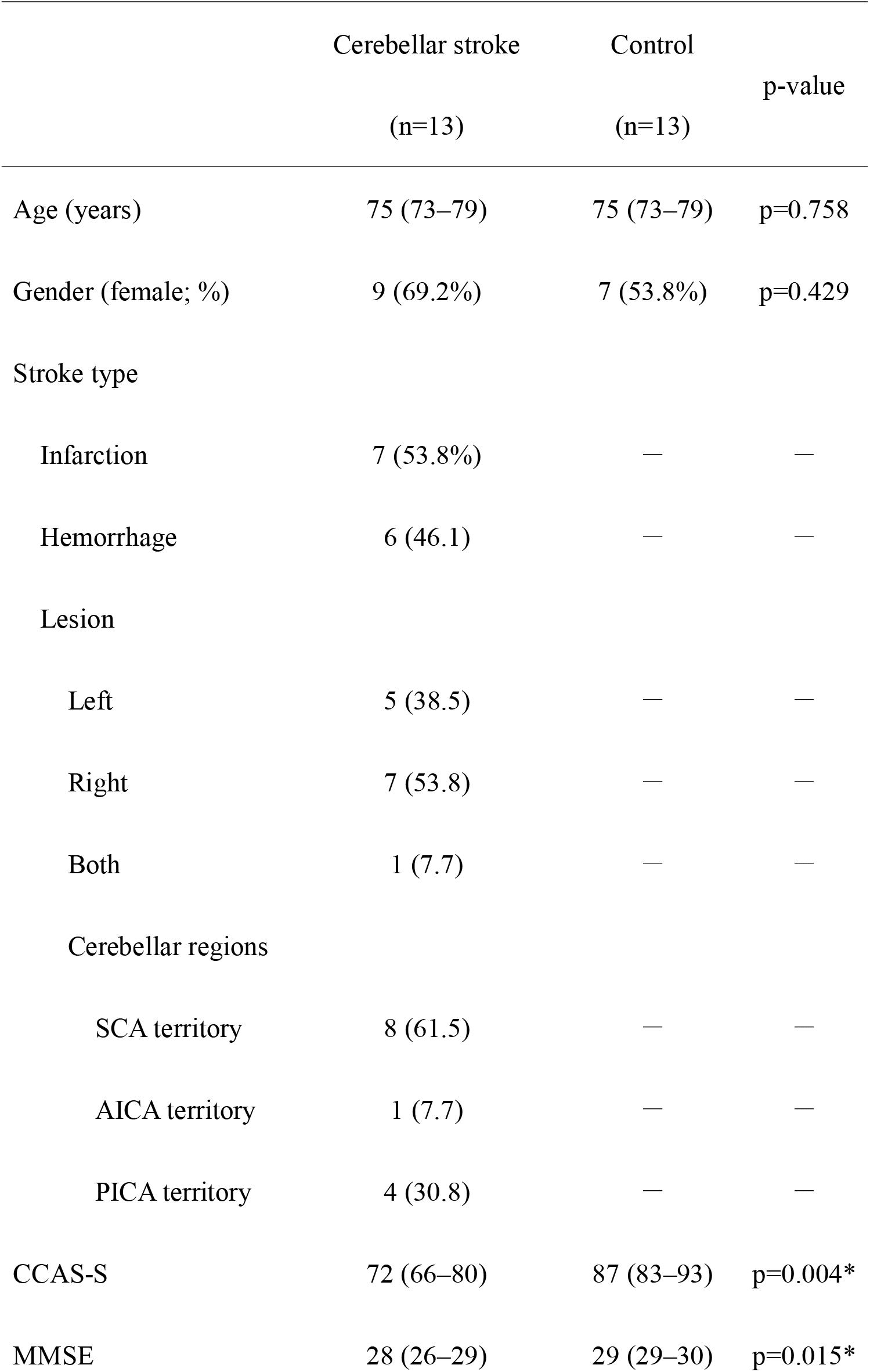
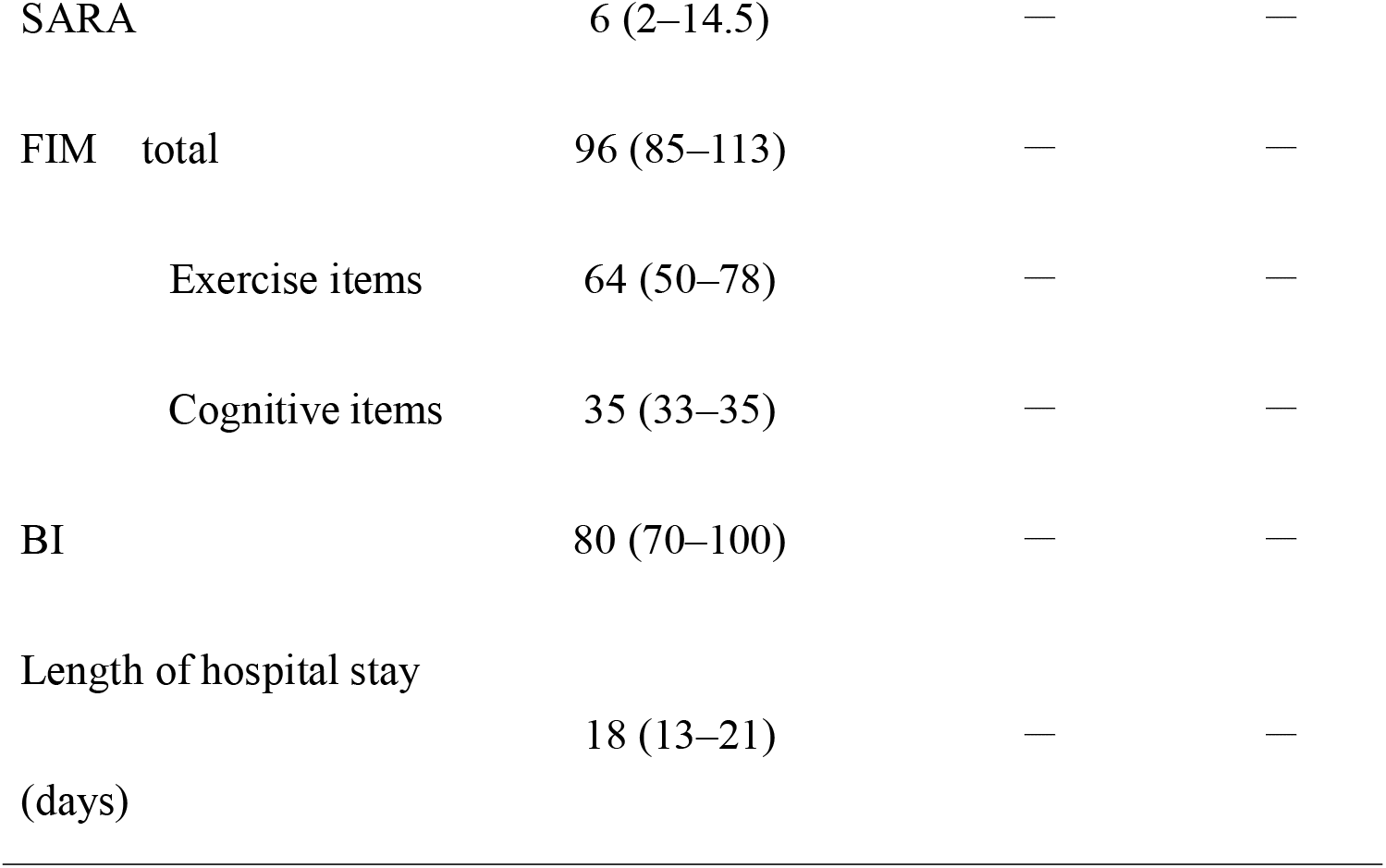
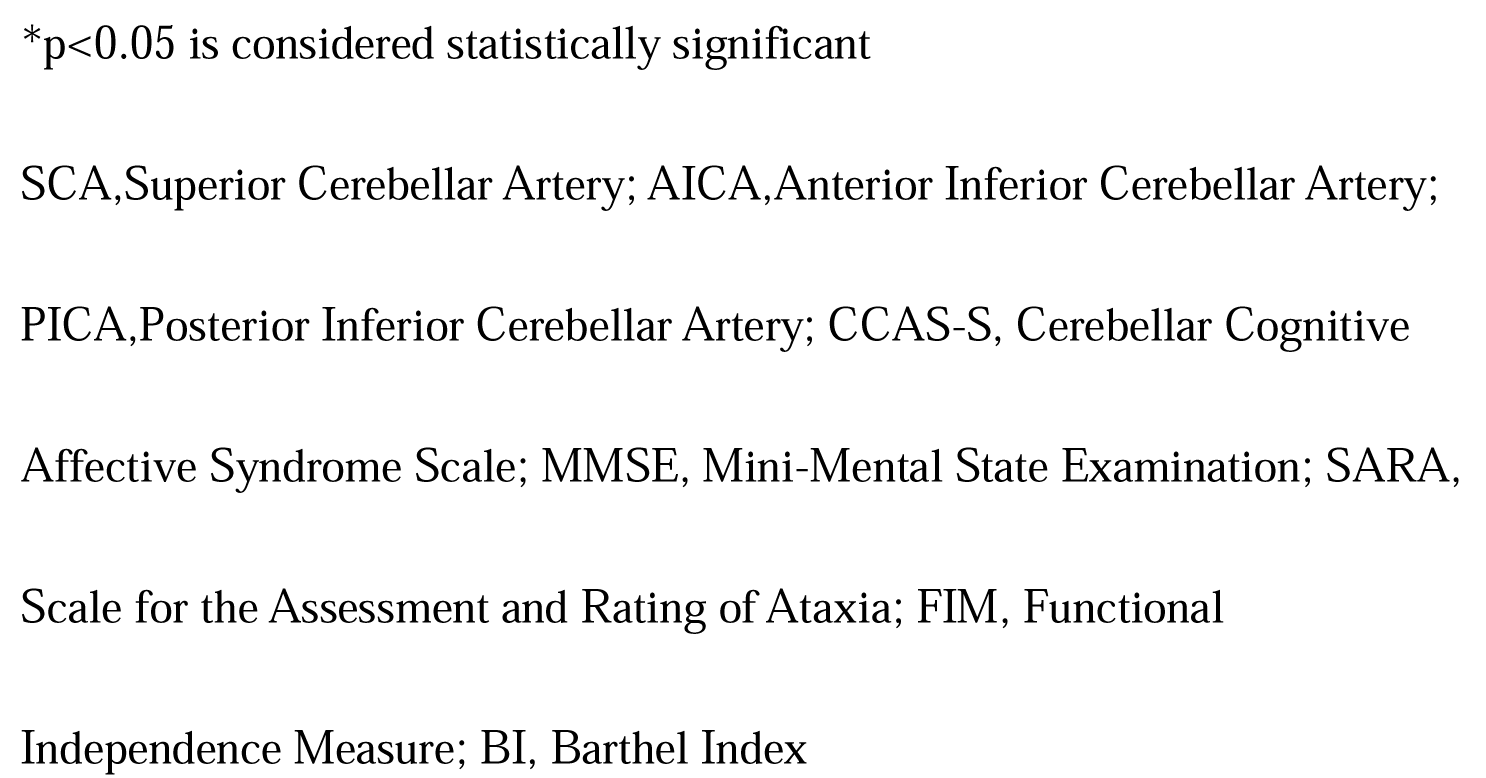
Patient characteristics of the study groups.

### CCAS in acute cerebellar stroke

Comparison of CCAS-S results with those of healthy controls is shown in Tables 1 and 2. All patients scored above the cutoff (23/24) on the MMSE; however, 11/13 (84.6%) scored below the cutoff (82 points) on the CCAS-S [8]. The median number of failed items per patient was four (IQR: 3–5). Consequently, 2, 1, and 10 patients were diagnosed with possible. probable, and definite CCAS, respectively. Frequently failed items were phonemic fluency (10 cases), verbal recall (nine cases), category switching (eight cases), semantic fluency (seven cases), and similarity (six cases). In contrast, the digit span forward, cube drawing, and digit span backward were less common (3, 2, and 1 case, respectively), and no patient was below the cutoff for Go-No-Go and Affect.

**Table 2.**
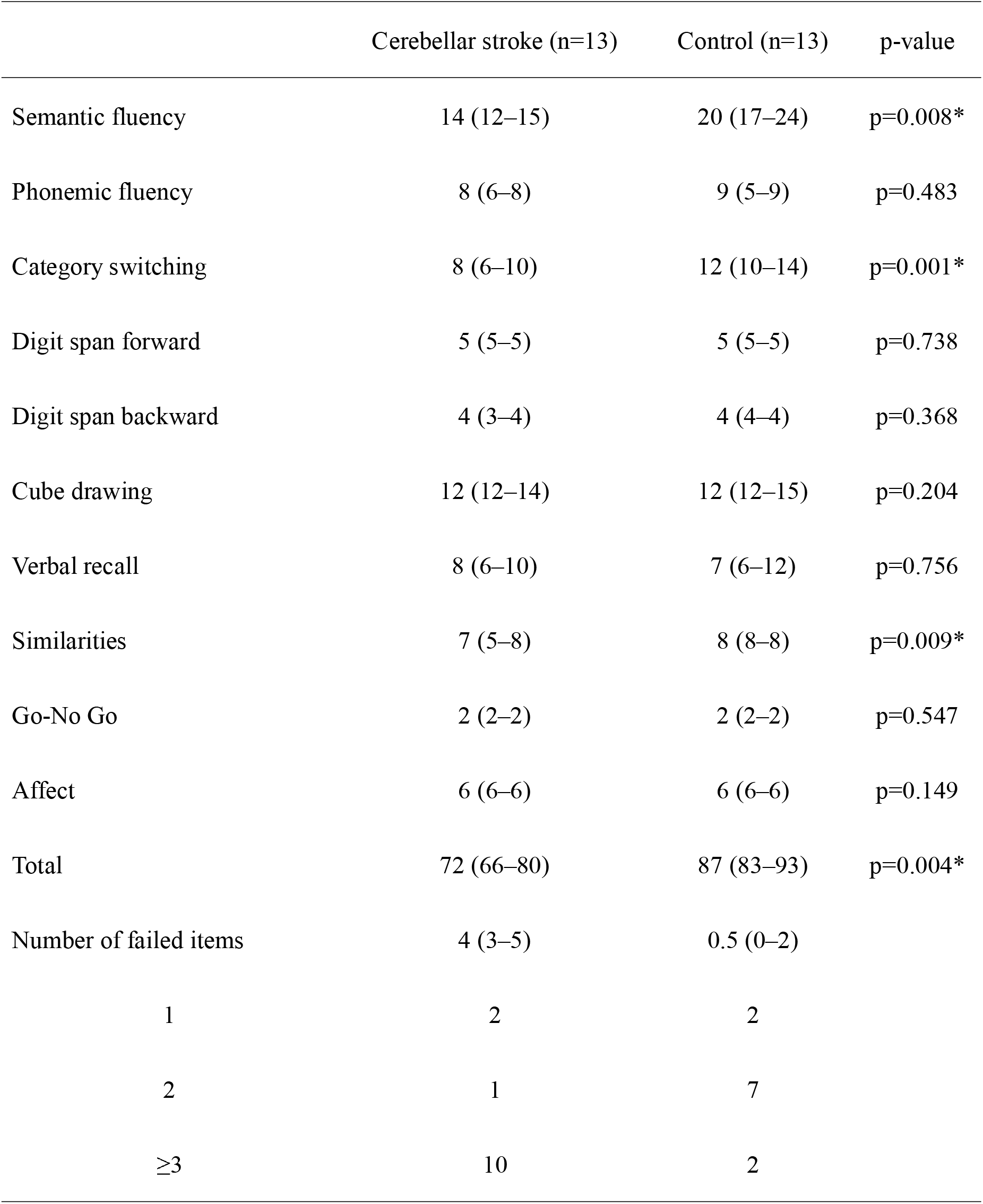
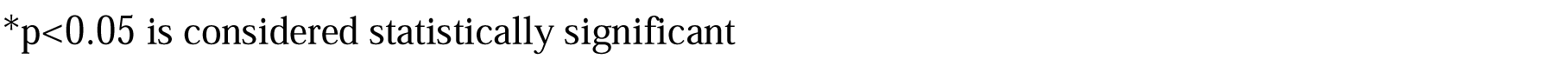
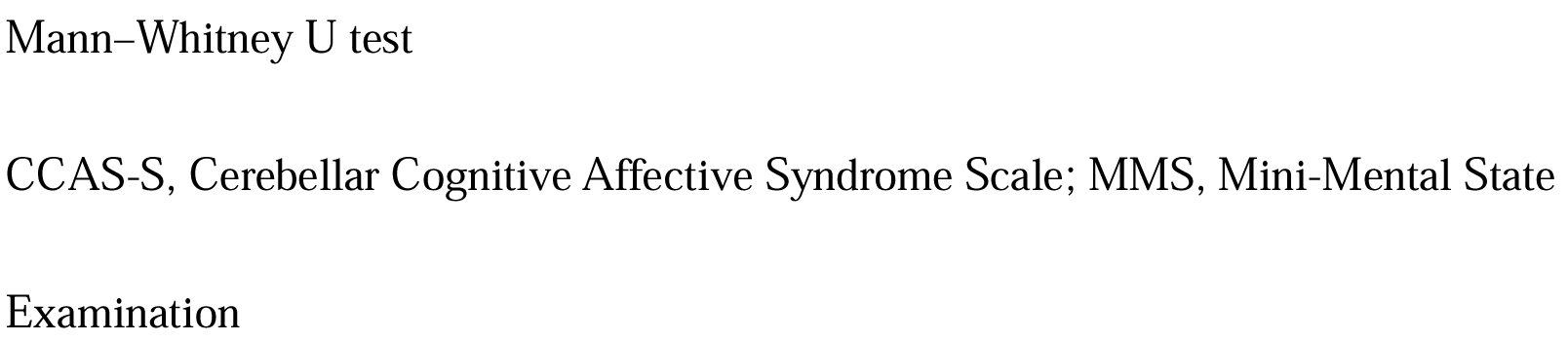
Comparison of CCAS-S in acute stroke and control groups.

A comparison of acute cerebellar stroke patients and healthy controls is shown in Table 2. In terms of CCAS-S scores, acute cerebellar stroke patients showed significantly lower values for total (p=0.004), semantic fluency (p=0.008), category switching (p=0.001), and similarities (p=0.009) (Figure 1) than controls. However, there were no significant differences in phonemic fluency (p=0.483) or verbal recall (p=0.756).

### CCAS and lesion location

Table 3 shows the lesion location and CCAS-S results for each patient (failed items, left: original cutoff; right: first quartile of controls as the cutoff). In four patients with total CCAS-S scores above 80, one and three patients were affected in vascular regions of SCA and non-SCA, respectively. Among the four patients with total CCAS-S scores between 70 and 80, three and one patients were affected in SCA and non-SCA, respectively. Among the five patients with total CCAS-S scores below 70, four and one patients were affected in SCA and non-SCA, respectively. Patients with lower total CCAS-S scores were more likely to have SCA lesions. In other words, SCA lesions tended to have lower total CCAS-S scores than the AICA/PICA lesions. In terms of cerebellar segmentation, SCA territory stroke caused more damage to the dentate nucleus (DN), whereas PICA territory stroke caused more damage to lobes VII and VIII.

**Table 3.**
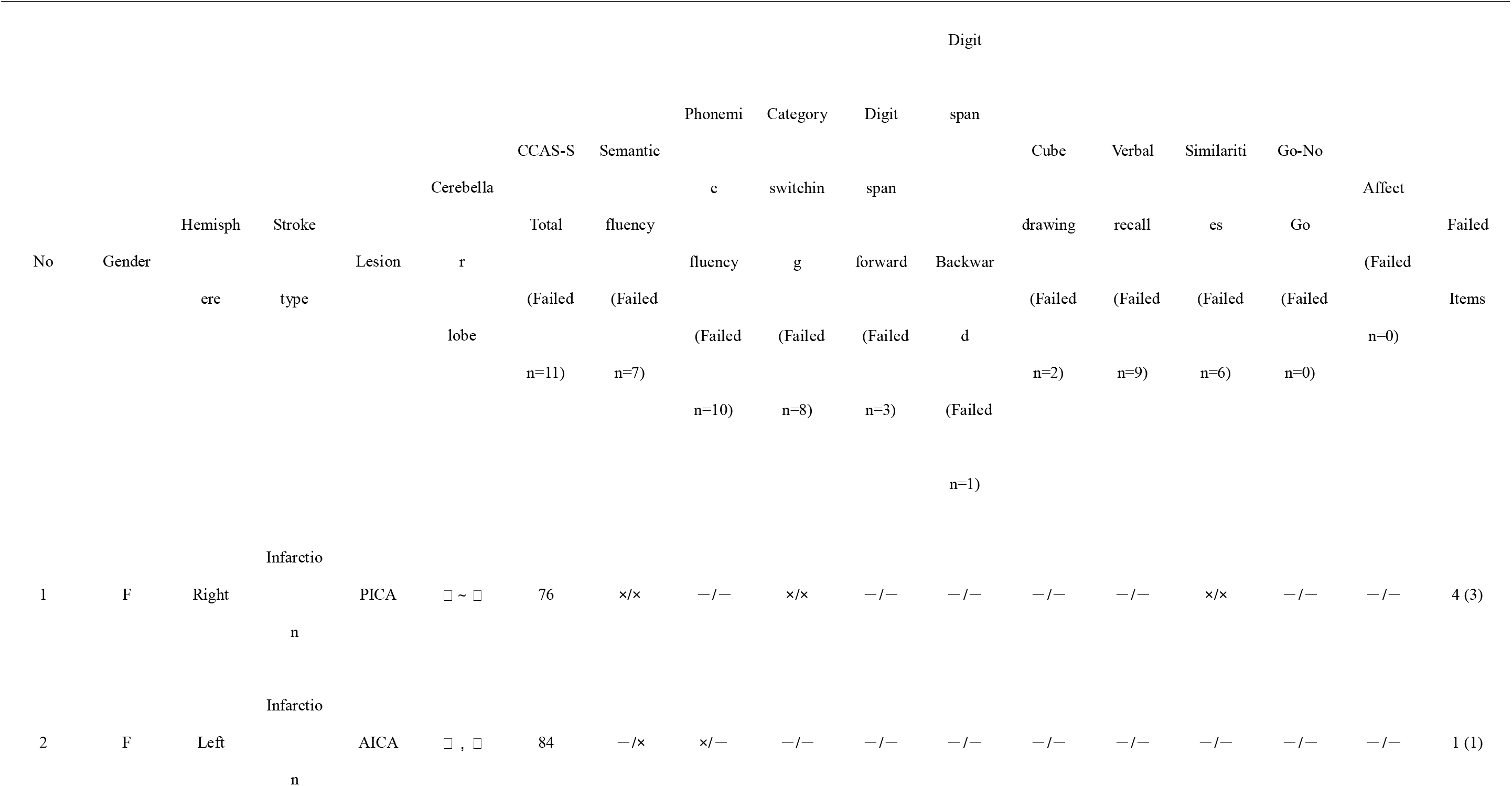
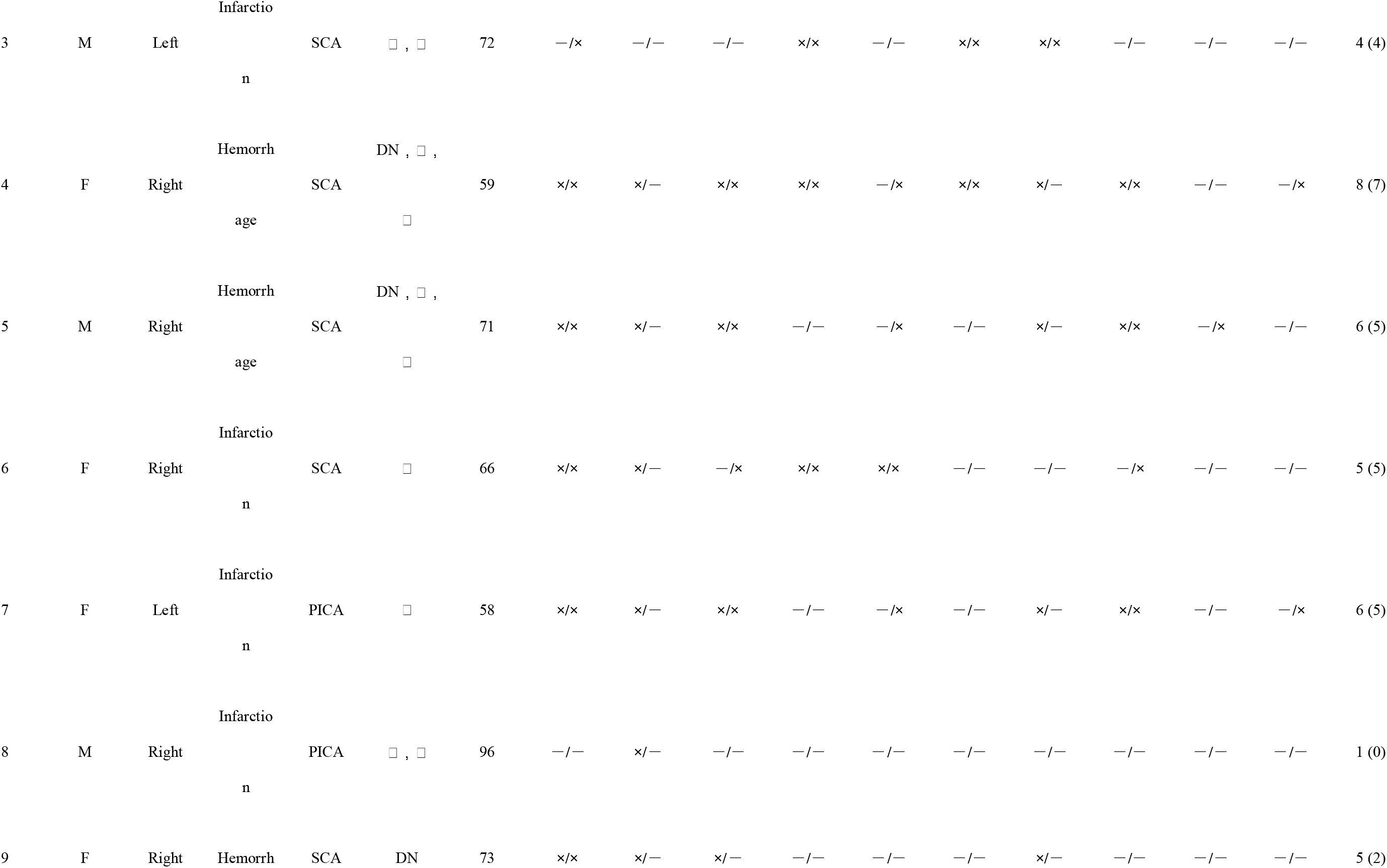
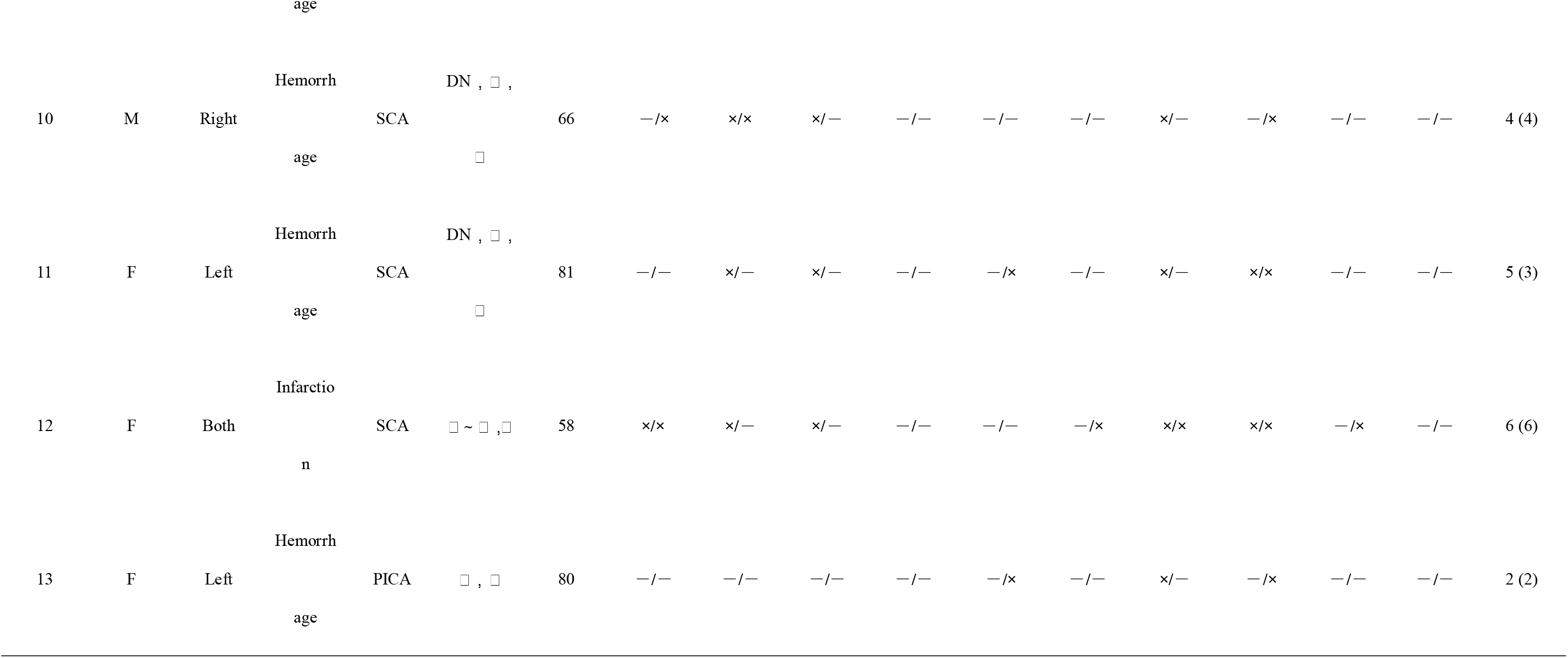
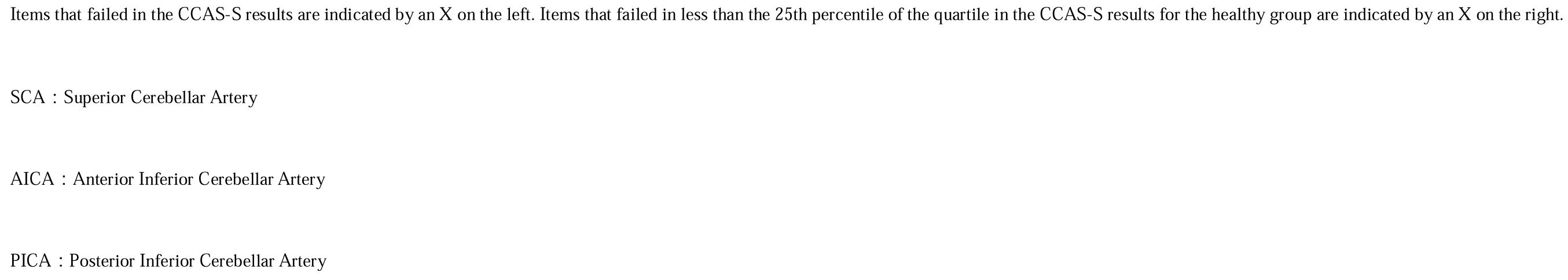
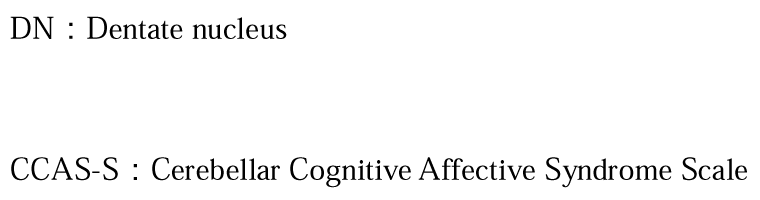
Patient characteristics and CCAS-S details.

### CCAS and outcome

Table 4 lists comparison of scores with the outcome. There were no significant differences in age (p=0.35), CCAS-S score (p=0.67), MMSE score (p=0.85), and hospitalization days (p=0.16) between the home discharge and transfer groups. However, the SARA (p=0.005), discharge FIM (p=0.003), and discharge BI (p=0.002) scores were significantly lower in the transfer group than in the home discharge group. There was no significant correlation between CCAS-S and SARA scores (r=0.233) (Figure 2).

**Table 4.**
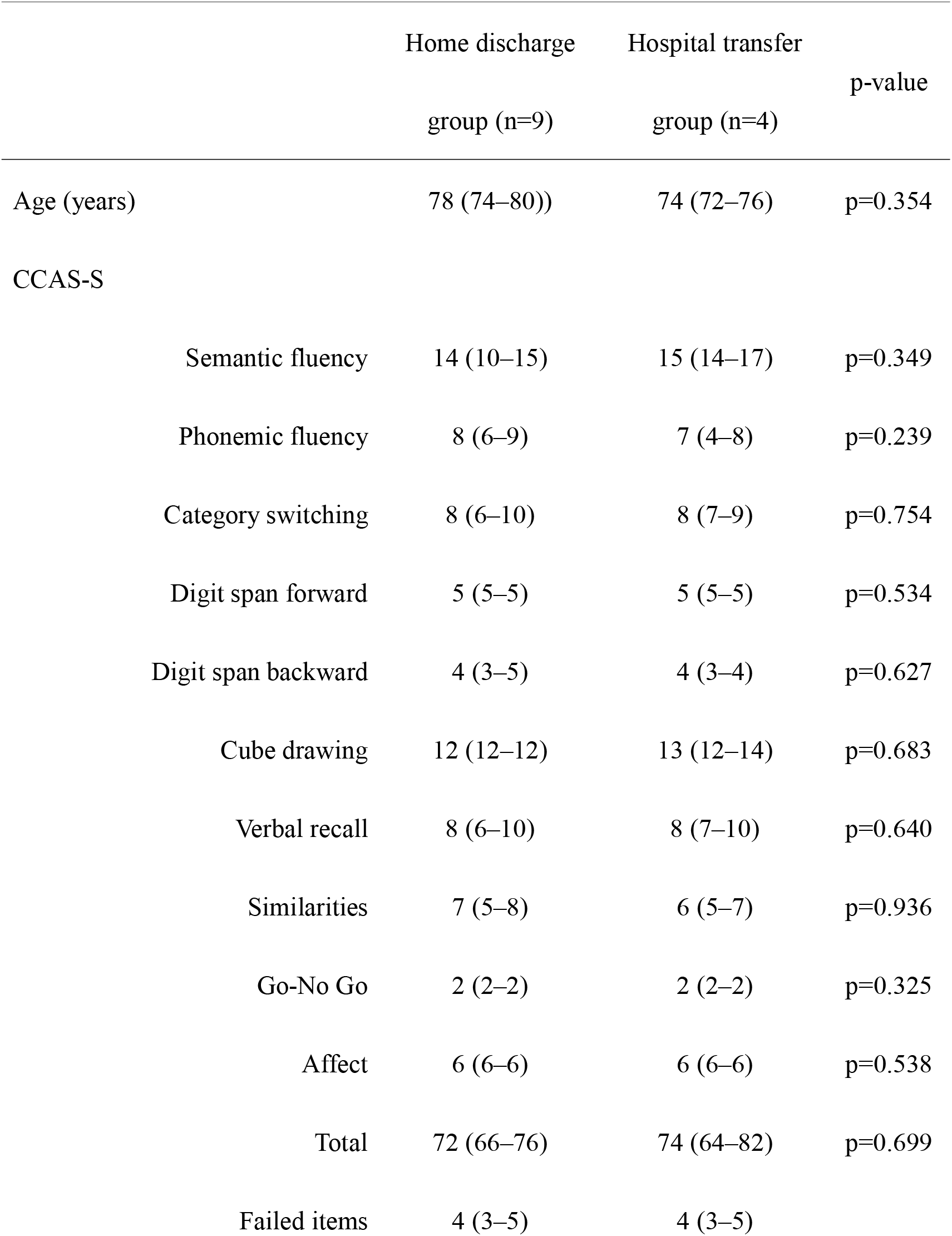
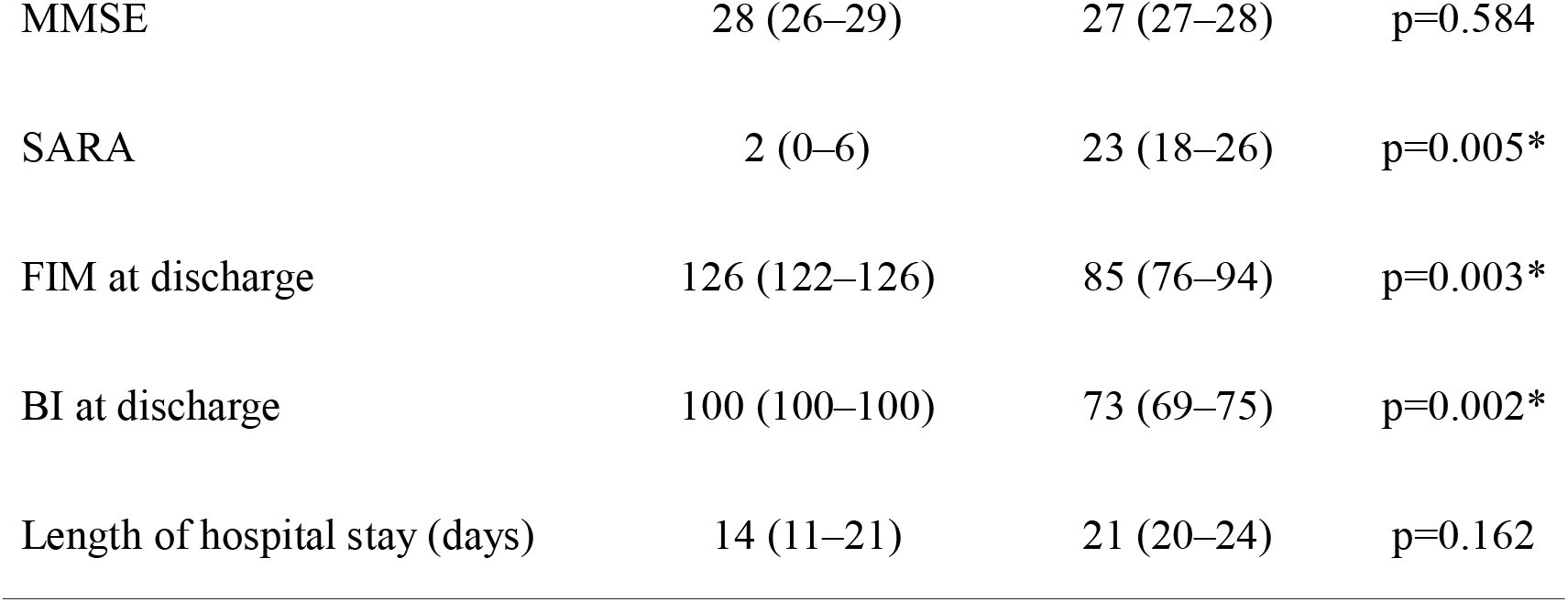
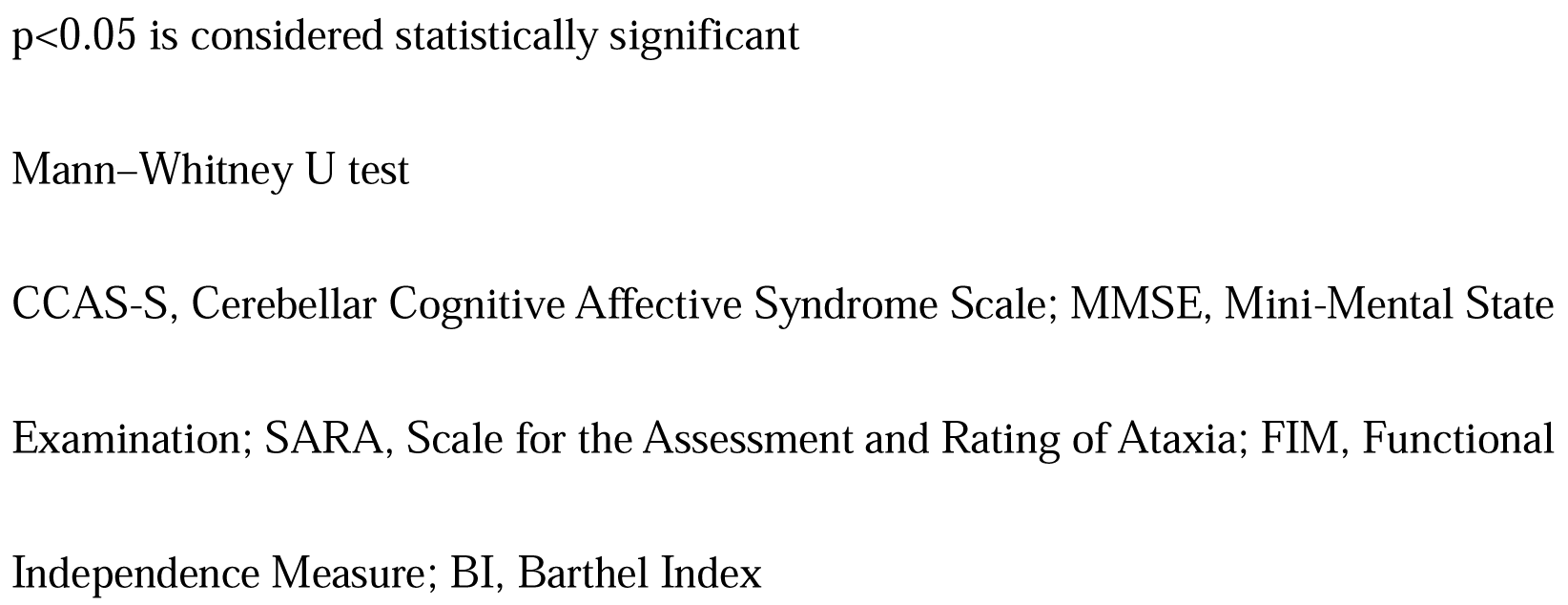
Impact of each item on the outcome.

## Discussion

This study investigated the characteristics of CCAS in patients experiencing their first acute cerebellar infarction. Based on the CCAS-S, 11/13 patients (84.6%) showed cognitive impairment due to cerebellar lesions; however, their MMSE score was within the normal range. The most frequently failed items in the CCAS-S were semantic fluency, category switching, and similarities compared with age-matched controls, and CCAS was more severe in SCA territory stroke than in AICA/PICA stroke. Despite the high frequency of CCAS in acute cerebellar stroke, it did not affect the outcome.

The CCAS-S is useful as a screening assessment for CCAS associated with cerebellar lesions [8]; however very few studies have used CCAS-S in patients with stroke, and only one study has reported the CCAS prevalence and used CCAS-S in acute stroke patients [11]. In a study conducted by Abderrakib et al., 25 patients with acute cerebellar stroke were evaluated with CCAS-S within one week of onset, and 1, 3, and 21 patients were classified as having possible, probable, and definite CCAS, respectively. In our study, 10/13 (76.9%) patients had definite CCAS, indicating that cognitive dysfunction is common in acute cerebellar stroke patients. Considering that the MMSE scores were normal in all our patients, the CCAS-S was useful in detecting CCAS.

Using the original cutoff value [8], the most frequently failed items were phonemic fluency (77%), verbal recall (69%), category switching (62%), semantic fluency (54%), and similarities (46%). However, this cutoff value has been criticized for not considering age and education [15]. Therefore, in this study, we compared these results with those of age-matched healthy controls. The CCAS-S items showing abnormalities included category switching, semantic fluency, and similarity. Low scores on the phonemic fluency and verbal recall items may be due to advanced age. In a previous study by Abderrakib et al. [11], category switching and backward digit span were the most commonly failed items, followed by phonemic fluency and verbal recall. The mean age was 64 years, which is younger than that in our study, and the results were based on the original cutoff value.

The advantage of the CCAS-S is that the severity and extent of CCAS can be compared between studies. In a study on spinocerebellar ataxia type 3, 22 patients with age of 51.9 ± 10.0 years were analyzed and the average total CCAS-S score was 81.9 ± 9.2 [16]. Regarding spinocerebellar ataxia type 2, 64 patients with a mean age of 48.77 ± 12.02 years showed a mean total CCAS-S score of 69.95 ± 13.48 [17]. Regarding chronic cerebellar stroke, 22 patients with age of 47.27 ± 13.10 years showed a mean total CCAS-S score of 88.18 ± 2.88 [18]. Considering the similar age of the patients in these three studies, CCAS might be most severe in spinocerebellar ataxia type 2 patients. This may be because the cerebellum and brainstem is affected in spinocerebellar ataxia type 2. Cognitive decline has been reported to occur even during the prodromal phase and is more pronounced in patients with severe motor activity in spinocerebellar ataxia type 2 [19]. The median total CCAS-S score for our patients was 72, which is similar to that of the spinocerebellar ataxia type 2 patients described above. Considering that the SARA score was higher in spinocerebellar ataxia type 2 patients than in our study, the similar total CCAS-S scores may be due to the advanced age of the patients.

In the above three studies, age- and sex-matched healthy participants were independently examined and compared with a patient group to determine which CCAS-S items were impaired. The spinocerebellar ataxia type 3 study revealed five failed items, including affect [16], and the spinocerebellar ataxia type 2 study found 10 failed items, including affect [17]. The chronic cerebellar stroke study found abnormalities in nine items, excluding affect [18]. In congruence, our study reported three failed items except for affect; degenerative diseases such as spinocerebellar ataxia type 2 and type 3 are more likely impair the affect item than cerebellar stroke. However, the use of independent healthy controls in each study may have introduced a bias; therefore, it would be desirable to establish a worldwide reference value for each age group.

The cerebellum is divided into 10 lobules based on the nomenclature introduced by Larsell [20], with lobules I–V as the anterior lobe, lobules VI–IX as the posterior lobe, and lobule X as the flocculonodular lobe. Lobule VII is further divided into lobules VIIa (crusts I and II) and VIIb. Previous studies have shown that the foci of CCAS are located in the posterior lobe (lobules VII and VIII, and mainly in the DN); specifically, in lobules VII and VIII for executive function, in VIIa and right lobule VIII for visuospatial cognition, and in the right VIIa and lobule IX for language function [21]. Other studies have indicated that the right posterolateral region is responsible for CCAS [18]. In the present study, the DN and lobes VI, VII, and VIII tended to be injured more frequently, supporting the results of previous studies.

The cerebrum and cerebellum are interconnected via neural networks. Strong connections have been established between the cerebellum and various regions including the prefrontal cortex, posterior parietal lobe, superior temporal lobe, and temporal lobe, indicating the cerebellar involvement in higher cognitive networks [22, 23]. The association between the cerebellum and brain can be categorized into motor and cognitive loop. In the context of CCAS, it is believed that the condition arises from a disruption in the cognitive loop, specifically involving the prefrontal cortex, bridge nucleus, cerebellar hemisphere/DN, and contralateral thalamus - dorsomedial nucleus of prefrontal cortex [24, 25]. A study reported that cerebellar lesions patients exhibited decreased cerebral blood flow and metabolism, particularly in premotor areas contralateral to the lesion [26]. Furthermore, a SPECT study revealed that, in cases where there was no damage to the brain regions under the tentorium, reduced blood flow in the contralateral medial frontal lobe resulting from a cerebellar lesion could potentially account for symptoms such as executive dysfunction, apathy, and impaired inhibition [27].

Although cognitive dysfunction and depression are known to affect outcomes after stroke [28–31], whether CCAS affects the outcomes of acute cerebellar stroke has not been examined. In our study, motor symptoms rather than CCAS were the factors affecting outcome; although CCAS was detected by the CCAS-S, the MMSE was normal in all patients, reflecting that cognitive impairment due to acute cerebellar stroke was not severe. In addition, studies of spinocerebellar ataxia type 2 and FRDA have reported an inverse association between CCAS-S and SARA. However, no significant correlation between CCAS-S and SARA was found in this study. This may be partly because, unlike patients with degenerative diseases, patients with cerebellar stroke experience only partial damage to the cerebellum.

There are a few limitations in this study. Firstly, the study was conducted at a single center and the number of patients was small. Secondly, the CCAS-S was administered in Japanese; thus, it could not be validated. Thirdly, the CCAS-S score of patients with acute cerebellar stroke was compared with healthy controls, as in previous studies.

## Conclusions

Despite the high frequency of CCAS in acute cerebellar stroke patients, it did not affect the outcome. In addition, the CCAS-S was useful in patients with acute cerebellar infarction, as most patients with acute cerebellar infarction in the present study had clear CCAS. Future studies should be conducted on a larger scale to examine the relationship with other neuropsychological tests. And since this study was conducted during a short acute period, the long-term prognosis of CCAS should also be investigated.

## Data Availability

The raw data supporting the conclusions of this article will be made available by the authors, without undue reservation.

## Non-standard abbreviations and acronyms

Cerebellar cognitive affective syndrome (CCAS)

Cerebellar cognitive affective syndrome-scale (CCAS-S)

Activities of daily life (ADL)

Mini-Mental State Examination (MMSE)

Scale for the Assessment and Rating of Ataxia (SARA)

Functional Independence measure (FIM)

Barthel Index (BI)

Friedreich’s ataxia (FRDA)

Superior, Anterior inferior, and Posterior inferior cerebellar arteries (SCA, AICA, and PICA)

## Acknowledgements

The authors thank Dr. Arakawa who contributed to the preparation of this study. We would like to thank Editage (www.editage.jp) for English language editing.

## References

1) Leiner HC, Leiner AL, Dow RS. Does the cerebellum contribute to mental skills? Behav Neurosci. 1986;100:443–454. https://doi.org/10.1037/0735-7044.100.4.443

2) Schmahmann JD, Sherman JC. The cerebellar cognitive affective syndrome. Brain. 1998;121:561–579. https://doi.org/10.1093/brain/121.4.561

3) Taskiran-Sag A, Uzuncakmak Uyanik H, Uyanik SA, Oztekin N. Prospective investigation of cerebellar cognitive affective syndrome in a previously non-demented population of acute cerebellar stroke. J Stroke Cerebrovasc Dis. 2020; 29:104923. https://doi.org/10.1016/j.jstrokecerebrovasdis.2020.104923

4) Ahmadian N, van Baarsen K, van Zandvoort M, Robe PA. The cerebellar cognitive affective syndrome-a meta-analysis. Cerebellum. 2019;18:941–950. https://doi.org/10.1007/s12311-019-01060-2

5) Omar D, Ryan T, Carson A, Bak TH, Torrens L, Whittle I. Clinical and methodological confounders in assessing the cerebellar cognitive affective syndrome in adult patients with posterior fossa tumours. Br J Neurosurg. 2014;28:755–764. https://doi.org/10.3109/02688697.2014.920487

6) Bolceková E, Mojzeš M, Van Tran Q, Kukal J, Ostrý S, Kulišťák P, Rusina R. Cognitive impairment in cerebellar lesions: a logit model based on neuropsychological testing. Cerebellum Ataxias. 2017;4:13. https://doi.org/10.1186/s40673-017-0071-9

7) Nerdal V, Gjestad E, Saltvedt I, Munthe-Kaas R, Ihle-Hansen H, Ryum T, Lydersen S, Grambaite R. The relationship of acute delirium with cognitive and psychiatric symptoms after stroke: a longitudinal study. BMC Neurol. 2022;22:234. https://doi.org/10.1186/s12883-022-02756-5

8) Hoche F, Guell X, Vangel MG, Sherman JC, Schmahmann JD. The cerebellar cognitive affective/Schmahmann syndrome scale. Brain. 2018;141:248–270. https://doi.org/10.1093/brain/awx317

9) Maas RPPWM, Killaars S, van de Warrenburg BPC, Schutter DJLG. The cerebellar cognitive affective syndrome scale reveals early neuropsychological deficits in SCA3 patients. J Neurol. 2021;268:3456–3466. https://doi.org/10.1007/s00415-021-10516-7

10) Naeije G, Rai M, Allaerts N, Sjogard M, De Tiège X, Pandolfo M. Cerebellar cognitive disorder parallels cerebellar motor symptoms in Friedreich ataxia. Ann Clin Transl Neurol. 2020;7:1050–1054. https://doi.org/10.1002/acn3.51079

11) Abderrakib A, Ligot N, Naeije G. Cerebellar cognitive affective syndrome after acute cerebellar stroke. Front Neurol. 2022;13:906293. https://doi.org/10.3389/fneur.2022.906293

12) Von Elm E, Altman DG, Egger M, Pocock SJ, Gøtzsche PC, Vandenbroucke JP, STROBE Initiative. The strengthening the reporting of observational studies in epidemiology (STROBE) statement: guidelines for reporting observational studies. Int J Surg. 2014;12:1495–1499. https://doi.org/10.1016/j.ijsu.2014.07.013

13) Sato K, Yabe I, Soma H, Yasui K, Ito M, Shimohata T, Onodera O, Nakashima K, Sobue G, Nishizawa M, et al. Reliability of the Japanese version of the Scale for the Assessment and Rating of Ataxia (SARA). Brain Nerve. 2009;61:591–595. https://doi.org/10.11477/mf.1416100488

14) Yamauchi K, Kumagae K, Goto K, Hagiwara R, Uchida Y, Harayama E, Tanaka S, Kuroyama S, Koyanagi Y, Arakawa S. Predictive validity of the scale for the assessment and rating of ataxia for medium-term functional status in acute ataxic stroke. J Stroke Cerebrovasc Dis. 2021;30:105631. https://doi.org/10.1016/j.jstrokecerebrovasdis.2021.105631

15) Thieme A, Röske S, Faber J, Sulzer P, Minnerop M, Elben S, Reetz K, Dogan I, Barkhoff M, Konczak J., et al. Reference values for the Cerebellar Cognitive Affective Syndrome Scale: age and education matter. Brain. 2021;144:e20. https://doi.org/10.1093/brain/awaa417

16) Maas RPPWM, Killaars S, van de Warrenburg BPC, Schutter DJLG. The cerebellar cognitive affective syndrome scale reveals early neuropsychological deficits in SCA3 patients. J Neurol. 2021;268:3456–3466. https://doi.org/10.1007/s00415-021-10516-7.

17) Rodríguez-Labrada R, Batista-Izquierdo A, González-Melix Z, Reynado-Cejas L, Vázquez-Mojena Y, Sanz YA, Canales-Ochoa N, González-Zaldívar Y, Dogan I, Reetz K, Velázquez-Pérez L. Cognitive decline is closely associated with ataxia severity in spinocerebellar ataxia Type 2: a validation study of the Schmahmann syndrome scale. Cerebellum. 2022;21:391–403. https://doi.org/10.1007/s12311-021-01305-z

18) Chirino-Pérez A, Marrufo-Meléndez OR, Muñoz-López JI, Hernandez-Castillo CR, Ramirez-Garcia G, Díaz R, Nuñez-Orozco L, Fernandez-Ruiz J. Mapping the cerebellar cognitive affective syndrome in patients with chronic cerebellar strokes. Cerebellum. 2022;21:208–218. https://doi.org/10.1007/s12311-021-01290-3

19) Rodríguez-Labrada R, Velázquez-Pérez L, Ortega-Sánchez R, Peña-Acosta A, Vázquez-Mojena Y, Canales-Ochoa N, Medrano-Montero J, Torres-Vega R, González-Zaldivar Y. Insights into cognitive decline in spinocerebellar ataxia type 2: a P300 event-related brain potential study. Cerebellum Ataxias. 2019;6:3. https://doi.org/10.1186/s40673-019-0097-2

20) Larsell O. Lobules of the mammalian and human cerebellum. Anat Rec. 1958;130:329–330

21) Stoodley CJ, MacMore JP, Makris N, Sherman JC, Schmahmann JD. Location of lesion determines motor vs. cognitive consequences in patients with cerebellar stroke. NeuroImage Clin. 2016;12:765–775. https://doi.org/10.1016/j.nicl.2016.10.013

22) Schmahmann JD. From movement to thought: anatomic substrates of the cerebellar contribution to cognitive processing. Hum Brain Mapp. 1996;4:174–198. https://doi.org/10.1002/(SICI)1097-0193(1996)4:3<174::AID-HBM3>3.0.CO;2-0

23) Schmahmann JD. The cerebrocerebellar system: anatomic substrates of the cerebellar contribution to cognition and emotion. Int Rev Psychiatry. 2001;13:247–260. https://doi.org/10.1080/09540260120082092

24) Krienen FM, Buckner RL. Segregated fronto-cerebellar circuits revealed by intrinsic functional connectivity. Cereb Cortex. 2009;19:2485–2497. https://doi.org/10.1093/cercor/bhp135

25) Lu X, Miyachi S, Takada M. Anatomical evidence for the involvement of medial cerebellar output from the interpositus nuclei in cognitive functions. Proc Natl Acad Sci U S A. 2012;109:18980–18984. https://doi.org/10.1073/pnas.1211168109

26) Broich K, Hartmann A, Biersack HJ, Horn R. Crossed cerebello-cerebral diaschisis in a patient with cerebellar infarction. Neurosci Lett. 1987;83:7–12. https://doi.org/10.1016/0304-3940(87)90207-2

27) Baillieux H, De Smet HJ, Dobbeleir A, Paquier PF, De Deyn PP, Mariën P. Cognitive and affective disturbances following focal cerebellar damage in adults: a neuropsychological and SPECT study. Cortex. 2010;46:869–879. https://doi.org/10.1016/j.cortex.2009.09.002

28) Park JH, Kim BJ, Bae HJ, Lee J, Lee J, Han MK, O Ky, Park SH, Kang Y, Yu KH, et al. Impact of post-stroke cognitive impairment with no dementia on health-related quality of life. J Stroke. 2013;15:49–56. https://doi.org/10.5853/jos.2013.15.1.49

29) Mijajlović MD, Pavlović A, Brainin M, Heiss WD, Quinn TJ, Ihle-Hansen HB, Hermann DM, Assayag EB, Richard E, Thiel A, et al. Post-stroke dementia – a comprehensive review. BMC Med. 2017;15:11. https://doi.org/10.1186/s12916-017-0779-7

30) Hadidi N, Treat-Jacobson DJ, Lindquist R. Poststroke depression and functional outcome: a critical review of literature. Heart Lung. 2009;38:151–162. https://doi.org/10.1016/j.hrtlng.2008.05.002

31) Taylor-Rowan M, Momoh O, Ayerbe L, Evans JJ, Stott DJ, Quinn TJ. Prevalence of pre-stroke depression and its association with post-stroke depression: a systematic review and meta-analysis. Psychol Med. 2019;49:685–696. https://doi.org/10.1017/S0033291718002003

